# Recovery Trends Show Greater Quadriceps Weakness After Patellar Tendon Versus Hamstring Autografts in ACL Reconstruction

**DOI:** 10.64898/2026.06.08.26355177

**Authors:** Benjiman Wilebski, Colin W. Bond, Benjamin C. Noonan, Sanford Health

## Abstract

**Context:** Although knee extensor and flexor strength deficits are well-documented after anterior cruciate ligament reconstruction, limited data exist characterizing how strength recovery evolves over time. Understanding the temporal patterns of recovery, and how they differ by autograft type, is critical for optimizing rehabilitation and return-to-sport decision-making.

**Objective:** To characterize temporal trends in knee extensor and flexor strength recovery during the first year post-ACLR and evaluate differences between patellar tendon and hamstring tendon autografts.

**Design:** Case series.

**Setting:** Sports physical therapy clinics within a large health system.

**Participants:** Five hundred three patients (17.8 ± 3.0 y) who underwent primary reconstruction with either patellar tendon or hamstring tendon autografts and completed a combined 730 return-to-sport tests within 12 months postoperatively.

**Main Outcome Measures:** Normalized peak isokinetic concentric knee extension and flexion torques for involved and uninvolved limbs, and normalized symmetry indices for knee extension and flexion strength.

**Results:** Knee extension strength on both limbs and extension strength symmetry improved over time. Patients with hamstring autografts demonstrated superior involved leg knee extension strength and better extension strength symmetry compared with those receiving patellar tendon autografts, although uninvolved leg strength was similar between autografts. Knee flexion strength on both limbs and flexion strength symmetry also improved over time. Patellar tendon autograft patients exhibited greater strength symmetry, despite no difference between autografts for flexion strength for the involved or uninvolved limb.

**Conclusions:** Autograft significantly influences muscle strength recovery following anterior cruciate ligament reconstruction. Hamstring tendon autografts are associated with superior recovery of knee extension strength and strength symmetry compared to patellar tendon autografts. These findings underscore the need for graft-specific rehabilitation strategies and earlier identification of patients at risk for delayed recovery.

## INTRODUCTION

Recovering thigh strength post-anterior cruciate ligament reconstruction (ACLR) is paramount to enhancing patient outcomes.^1-3^ This recovery is intricately linked to the choice of autograft, including bone-patellar tendon-bone (BTB) and hamstring tendon (HAM) autografts, which fundamentally influence the function of the knee extensors (KE) and flexors (KF).^4-6^ Persistent KE and KF muscle weakness following ACLR has been linked to suboptimal outcomes.^1,2,7,8^ Such muscle weakness may comprise the knee joint’s capacity to attenuate joint and soft tissue forces and maintain active stabilization, potentially undermining movement strategies, reducing neuromuscular performance, and elevating the risk of ACL reinjury.^1,3,9-14^

Sports medicine providers frequently measure KE and KF muscle strength using a dynamometer, primarily through concentric isometric or isokinetic contractions, to evaluate readiness for a return to sport (RTS) post-ACLR. ^15,16^ Metrics such as instantaneous peak torque, adjusted for body mass, serve as an indicator of muscle strength.^17^ The normalized symmetry index (NSI), or another comparable symmetry based metric that compares the strength of the involved leg to the uninvolved leg, provides additional information and achieving a degree of KE muscle strength symmetry is often a primary criterion in the RTS decision-making process.^18,19^ These metrics offer unique insights, underlining the complexity of muscle strength assessment and suggesting that a comprehensive approach is essential for a holistic characterization of muscle strength recovery.^1,16,20^

Improving our understanding of how time post-ACLR and autograft influence muscle strength recovery is critical. These insights can aid providers in the characterization of a patient’s muscle strength recovery post-ACLR, which can in turn facilitate the development of customized rehabilitation plans and enable earlier identification of patients at risk for suboptimal strength recovery, well before reaching the desired RTS time.^21^ Accordingly, the purpose of this study was to assess the effects of autograft and time post-ACLR on KE and KF muscle strength in adolescent and young-adult patients up to 12 months post-ACLR. Our hypothesis was that KE NSI would become more symmetrical over time post-ACLR, with HAM patients exhibiting more symmetrical KE NSI compared to BTB patients.

## METHODS

### Subjects

The Sanford Health Institutional Review Board granted the retrospective analysis an exemption (review number: 2390). Beginning in 2012, post-ACLR RTS test data from patients who completed RTS tests at our health system’s sports physical therapy clinics were added to our database. Anonymous data from this database was provided by an honest broker on January 23^rd^, 2025. At this time, the full database contained data from 1,125 ACLR patients. Patients who underwent primary ACLR and received BTB or HAM who were greater than 12-but less than 30-years-old at the time of ACLR were included in this analysis if they had one or more post-ACLR RTS tests between 0- and 12-months post-ACLR, which included a KE and KF muscle strength test. Patients were included regardless of articular cartilage or menisci pathologies treated at the time of ACLR. Patients were excluded from the analysis if they had a history of significant lower-extremity injury prior to the present ACL injury including a previous ACLR, an allograft or autograft supplemented with allograft, post-ACLR complications that required secondary surgery for the present ACL injury, another knee ligament repaired or reconstructed in addition to the ACL, an autograft harvested from the contralateral limb, a quadriceps tendon autograft, or waited longer than 1-year between injury and ACLR. After exclusions, 503 ACLR patients with 730 post-ACLR RTS tests representing 33 unique surgeons were included (Table 1).

**Table 1.**
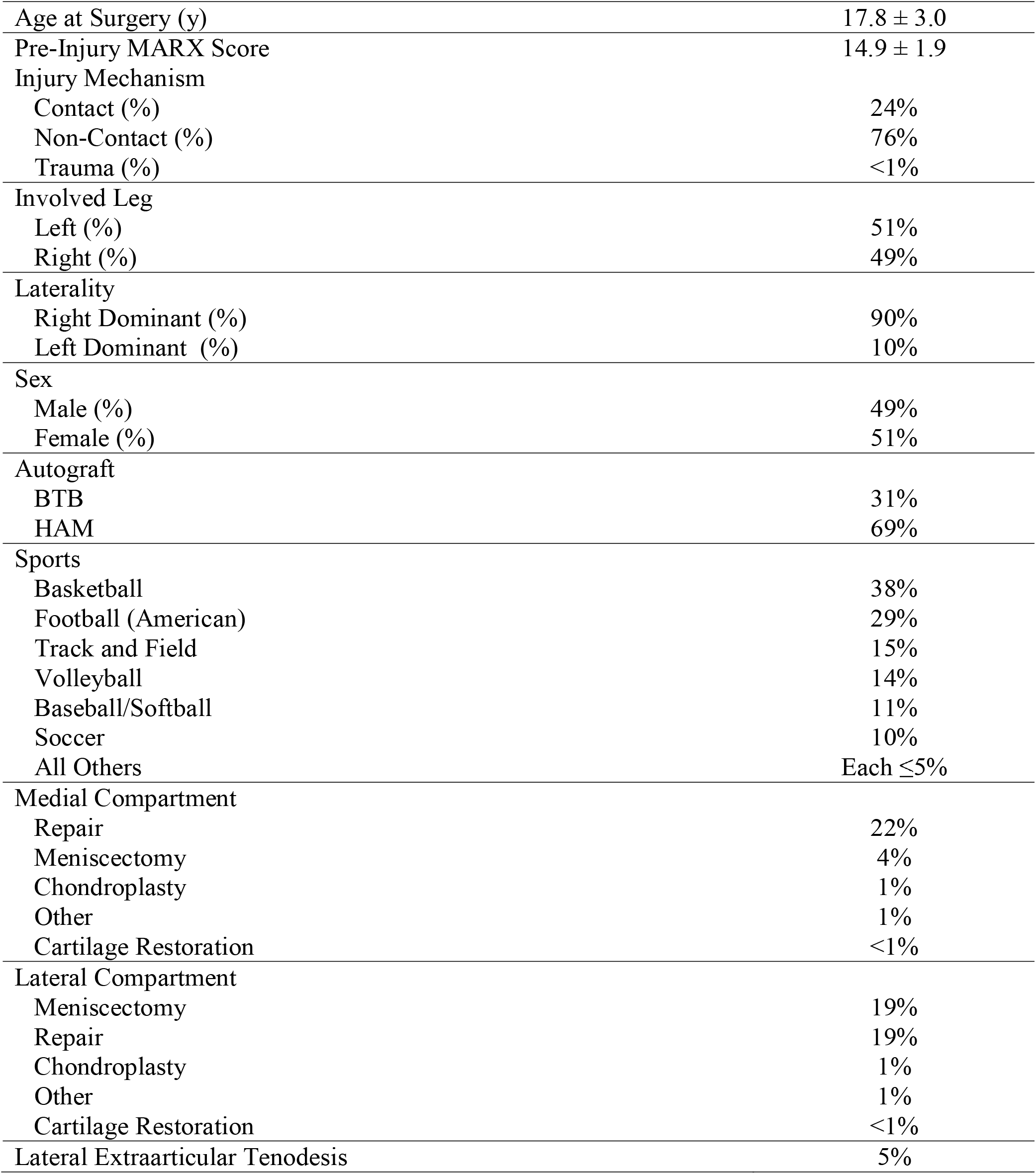
Demographic data. Data presented as mean ± standard deviation or percentage.

### Procedures

Our health system follows a criterion-based, individualized rehabilitation protocol post-ACLR. Early, progressive weight-bearing is initiated immediately post-operatively in the absence of concomitant procedures, with brace use and progression guided by quadriceps control and joint stability. The program advances through staged phases emphasizing restoration of range of motion, neuromuscular control, strength, and movement quality, with one or more RTS tests post-ACLR to guide rehabilitation and augment RTS decision-making. Tests administered during the post-ACLR RTS test include, but are not limited to, lower-extremity strength measured as instantaneous peak torque using a dynamometer. Different dynamometers were used at our sports physical therapy clinics; although, previous literature has indicated that dynamometers from different manufacturers provide similar results.^22^ Additionally, previous literature as suggested that measures of peak torque have high test-retest and intra-rater reliability.^23,24^ Patients were positioned in the dynamometer so that their knee axis of rotation was aligned with the dynamometer’s axis of rotation and their shank hung parallel with the dynamometer’s moment arm. The dynamometer’s pad was secured to the anterior aspect of the patient’s distal shank approximately 5 cm proximal to the ankle. Patients were then secured in the dynamometer using a strap around the test leg’s thigh and straps around the torso.

Patients completed concentric KE and KF testing on the uninvolved leg first followed by the involved leg at 60 deg·s^-1^, which is the most frequently utilized angular velocity during RTS tests ^16^. Torque was assessed through a range of 0° (considered full extension) to 90° of flexion.

Prior to the test set on each leg, patients were given a familiarization set of three to five repetitions. Patients were then given up to 30 seconds of rest before the test set. For the test set, patients performed five repetitions at a maximal effort. One repetition included a concentric KE action immediately followed by a concentric KF action. The peak instantaneous torque (lbf-ft) recorded during the test set for KE and KF, respectively, was initially printed on paper reports and then was subsequently transferred to a digital clinical data repository managed using REDCap.

### Data Preparation

The peak instantaneous KE and KF torques recorded during the test set were normalized to the patient’s body weight (lbf-ft·lb^-1^) for the involved (KE INV and KF INV) and uninvolved (KE UNINV and KF UNINV) leg. The NSI was calculated for KE (KE NSI) and KF (KF NSI).^18^ The NSI can range from 0% (full symmetry) to ±100% (full asymmetry), with negative values indicating an INV deficit and positive values indicating an UNINV deficit.

### Statistical Analyses

We robustly checked for outliers by computing the multivariate Mahalanobis distances using the observed KE INV, KF INV, KE UNINV, KF UNINV, and time post-ACLR. Five outliers were identified and subsequently removed. The continuous dependent variables were KE INV, KF INV, KE UNINV, KF UNINV, KE NSI, and KF NSI. The continuous independent variables were time post-ACLR measured in months and the categorical independent variable was autograft. The co-occurrence of a meniscal repair or cartilage restoration procedure, which would necessitate a longer period of limited weight-bearing post-ACLR, was considered as a covariate to control for the potential effects on KE and KF muscle strength post-ACLR. A mixed effects model was used to explore the relation between each dependent variable and the interaction between autograft and time post-ACLR with random intercepts by patient. The within-subject correlation over time post-ACLR was modeled using a continuous autoregressive structure. Variance was modeled as a function of time post-ACLR and autograft. We robustly assessed the model’s standardized residuals for normality. Subsequent analysis of variance (ANOVA) was used to evaluate the effects of time post-ACLR and autograft on the dependent variables. Statistical significance was set at p < .05. Data were analyzed using R (version 4.4.1).

We conducted an empirical, resampling-based power analysis using our full dataset and model. Stratified Monte Carlo resampling by autograft was performed across a range of training sample proportions with 1,000 resamples per proportion. For each resample, the model was refit and statistical significance was assessed for the fixed effects of time post-ACLR and autograft on the primary dependent variable KE NSI, aligning with our hypothesis. The analysis indicated that a sample size of approximately 113 patients—comprising 78 HAM and 35 BTB patients— was required to achieve 80% power to detect the effects of time post-ACLR and autograft at a significance level of 0.05.

## RESULTS

### Knee Extension

For KE INV, there were significant effects of time post-ACLR (4.1 [3.5, 4.7]; p < .001) and autograft (20.1 [13.9, 26.4]; p < .001) as KE INV increased with time post-ACLR and HAM patients had greater KE INV than BTB patients, but there was no evidence of an interaction effect (-.7 [-1.7, .2]; p = .115) (Figure 1). For KE UNINV, there was a significant effect of time post-ACLR (1.2 [.4, 1.9]; p = .002) as KE UNINV increased with time post-ACLR, but there was no evidence of an effect of autograft (4.6 [-2.6, 11.8]; p = .208) or interaction (-.5 [-1.5, .5]; p = .307) (Figure 2). Finally, for KE NSI, there were significant effects of time post-ACLR (3.4 [2.6, 4.2]; p < .001) and autograft (21.4 [14.2, 28.5]; p < .001) as KE NSI increased with time post-ACLR and HAM patients had greater KE NSI than BTB patients, but there was no evidence of an interaction effect (-.7 [-1.7, .4]; p = .194) (Figure 3).

**Figure 1.**
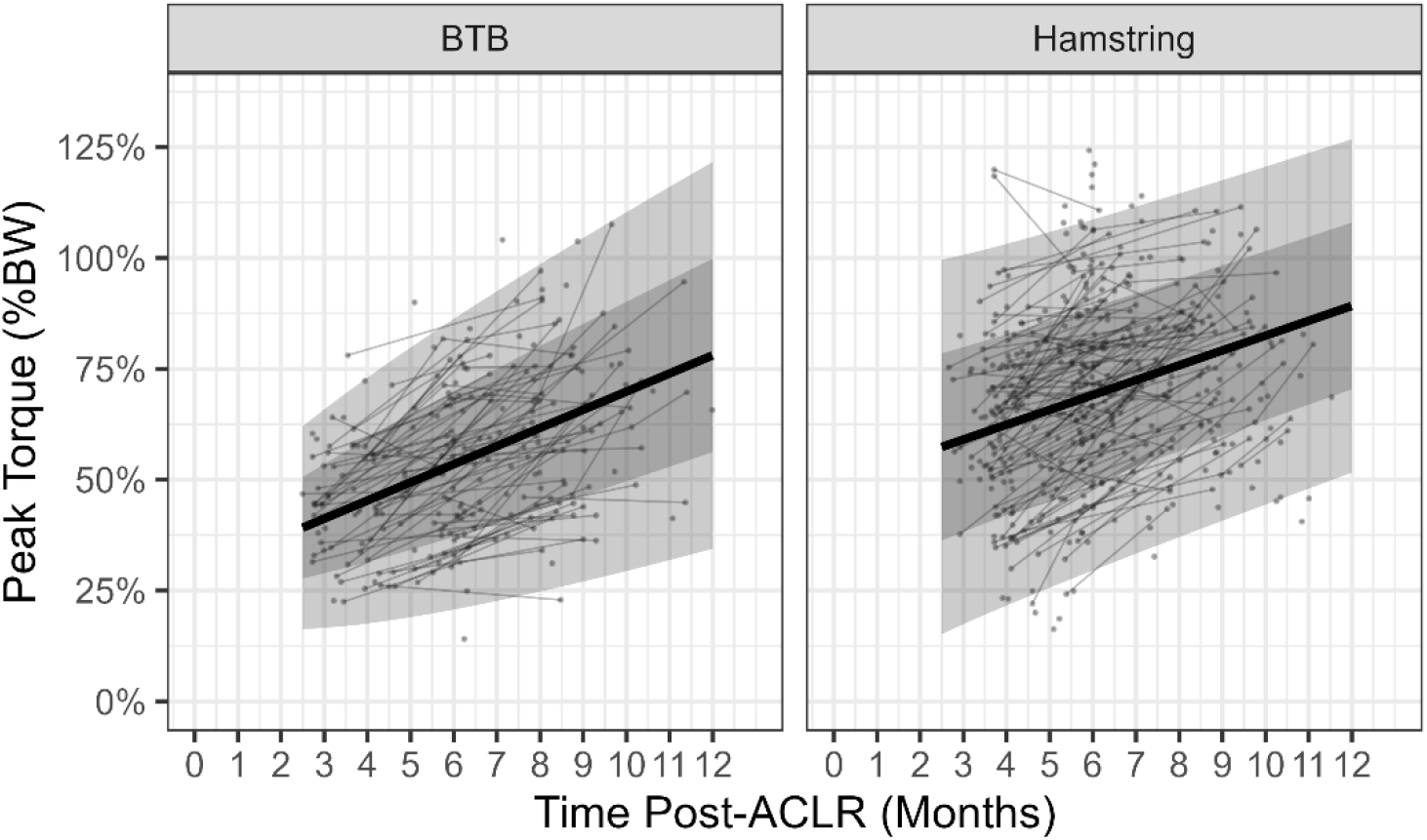
Involved leg normalized knee extension peak torque (% body weight) as a function of time post-anterior cruciate ligament reconstruction (ACLR) and autograft for bone-patella tendon-bone (BTB) and hamstring autografts. Thin dots and lines: individual patients; thick line: modeled mean; dark grey ribbon: one standard deviation; light grey ribbon: two standard deviations.

**Figure 2.**
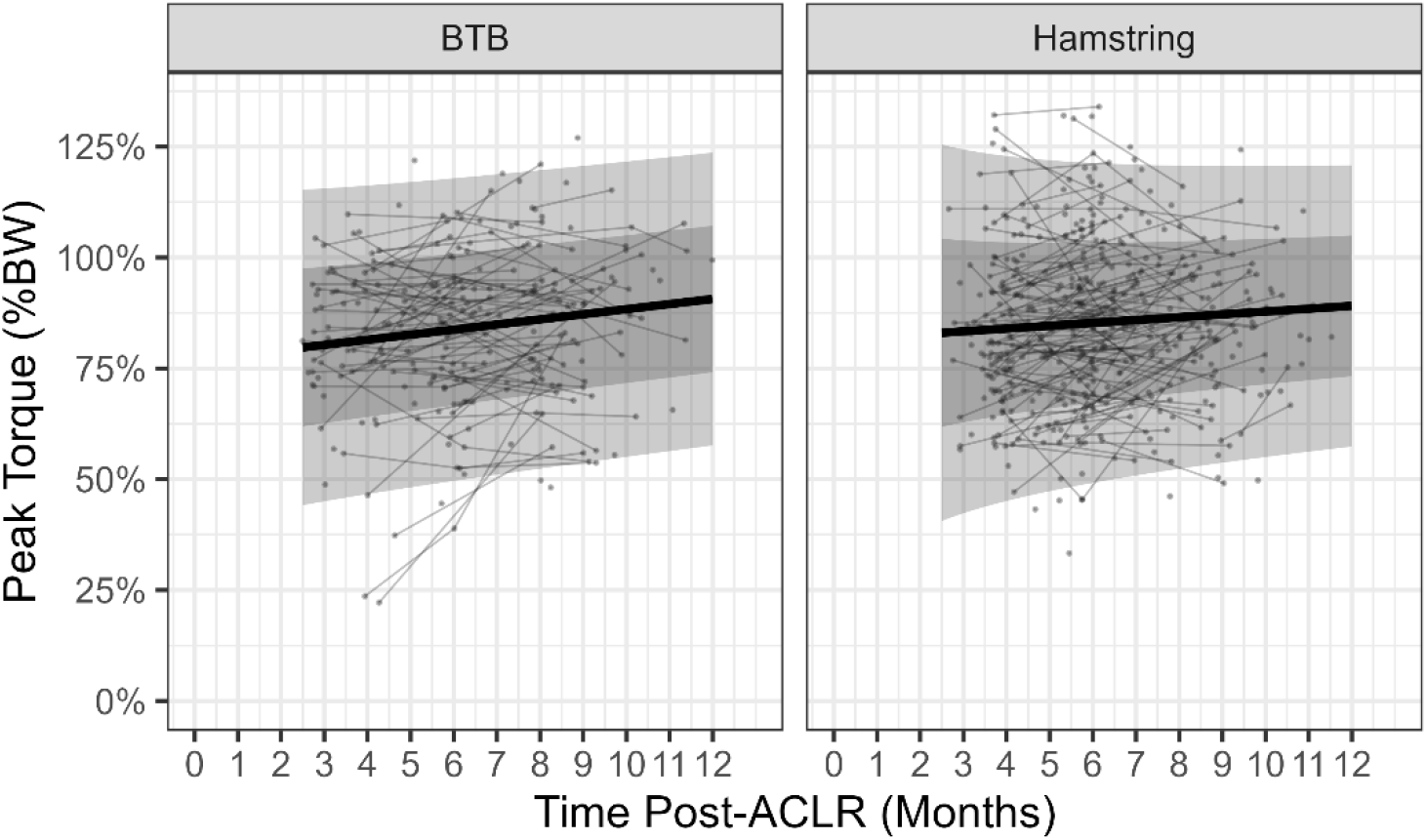
Uninvolved leg normalized knee extension peak torque (% body weight) as a function of time post-anterior cruciate ligament reconstruction (ACLR) and autograft for bone-patella tendon-bone (BTB) and hamstring autografts. Thin dots and lines: individual patients; thick line: modeled mean; dark grey ribbon: one standard deviation; light grey ribbon: two standard deviations.

**Figure 3.**
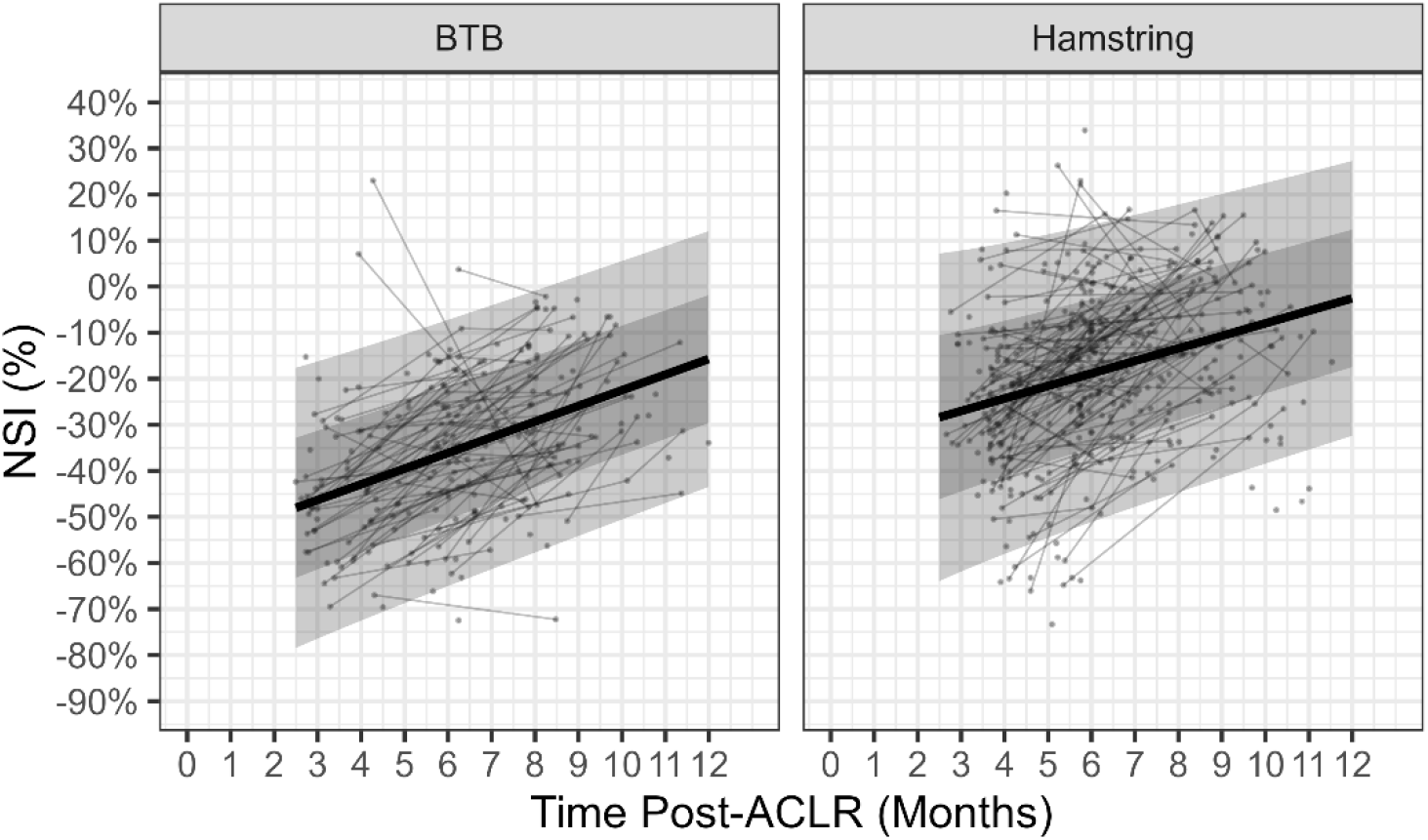
Normalized symmetry index (NSI) for knee extension strength as a function of time post-anterior cruciate ligament reconstruction (ACLR) and autograft for bone-patella tendon-bone (BTB) and hamstring autografts. Thin dots and lines: individual patients; thick line: modeled mean; dark grey ribbon: one standard deviation; light grey ribbon: two standard deviations.

### Knee Flexion

For KF INV, there was a significant effect of time post-ACLR (1.8 [1.4, 2.1]; p < .001) as KF INV increased with time post-ACLR, but there was no evidence of an effect of autograft (-5.1 [-1.6, 2.0]; p = .379) or interaction (-.3 [-.7, .2]; p = .296) (Figure 4). For KF UNINV, there was a significant effect of time post-ACLR (1.1 [.6, 1.5]; p < .001) as KF UNINV increased with time post-ACLR, but there was no evidence of an effect of autograft (3.1 [-1.1, 7.3]; p = .142) or interaction (-.5 [-1.2, .1]; p = .079) (Figure 5). Finally, for KF NSI, there was a significant effect of time post ACLR (1.2 [.5, 1.9]; p < .001) and autograft (-11.4 [-18.0, -4.7]; p < .001) as KF NSI increased with time post-ACLR and BTB patients has greater KF NSI than HAM patients, but there was no evidence of an interaction effect (.8 [-.2, 1.7]; p = .125) (Figure 6).

**Figure 4.**
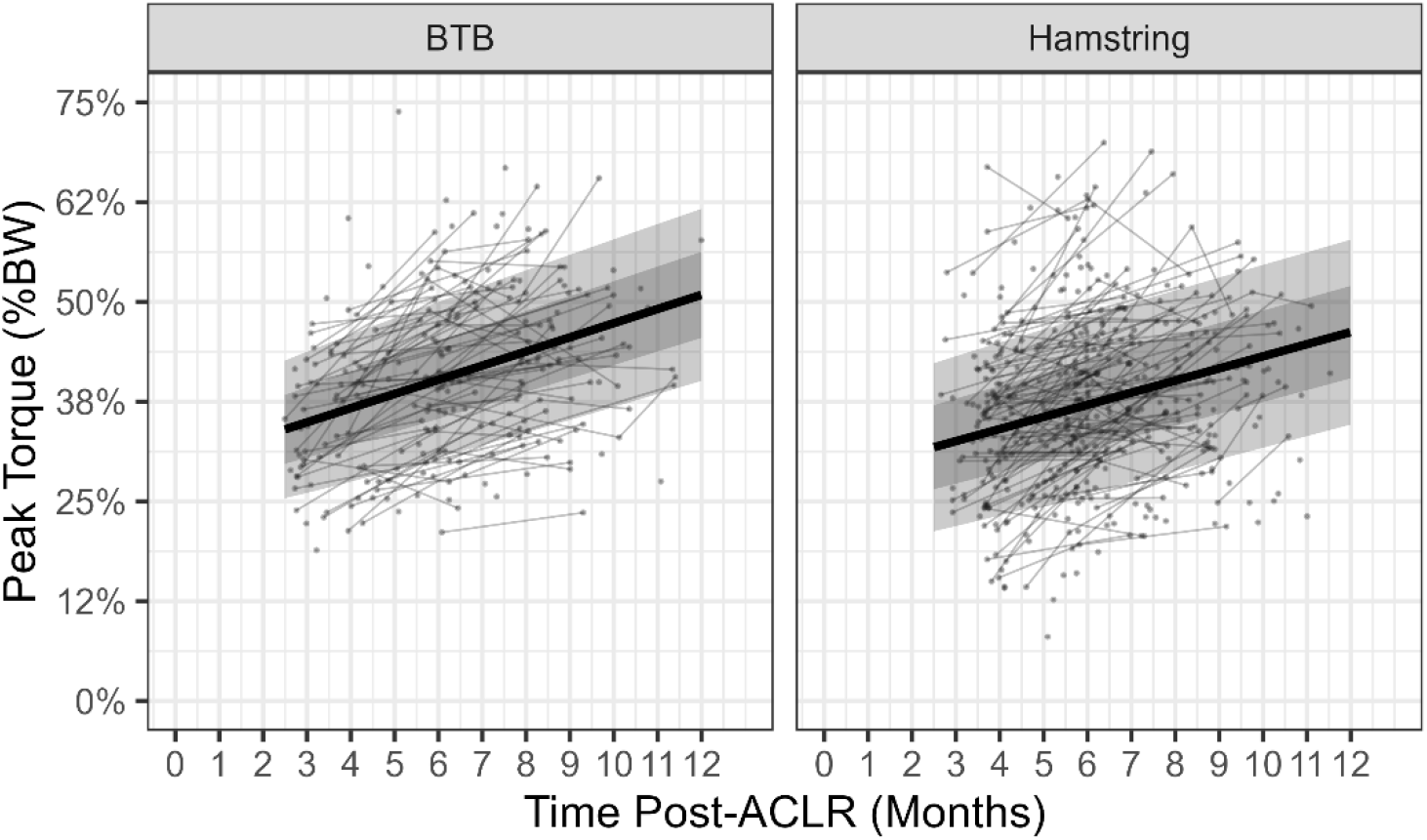
Involved leg normalized knee flexion peak torque (% body weight) as a function of time post-anterior cruciate ligament reconstruction (ACLR) and autograft for bone-patella tendon-bone (BTB) and hamstring autografts. Thin dots and lines: individual patients; thick line: modeled mean; dark grey ribbon: one standard deviation; light grey ribbon: two standard deviations.

**Figure 5.**
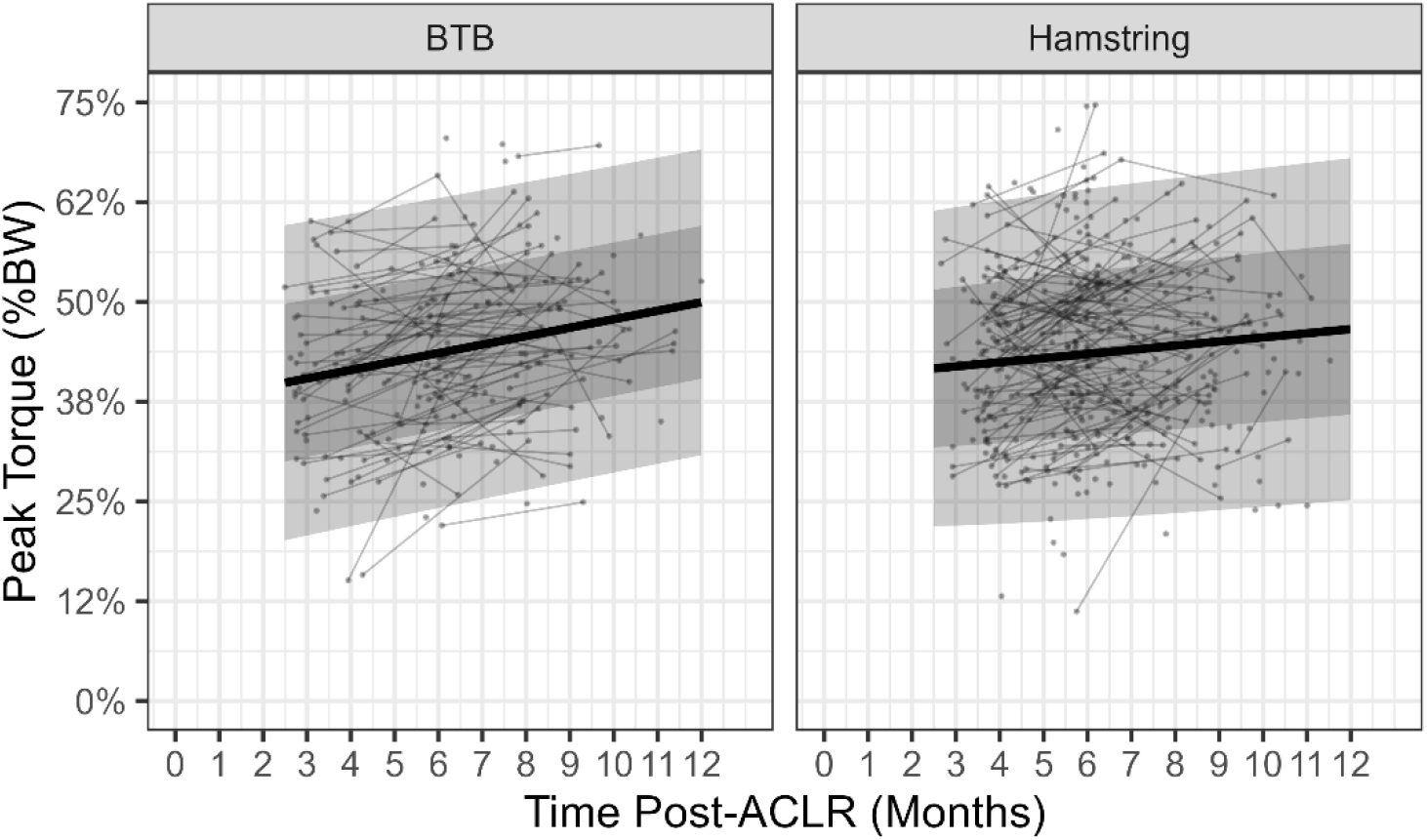
Uninvolved leg normalized knee flexion peak torque (% body weight) as a function of time post-anterior cruciate ligament reconstruction (ACLR) and autograft for bone-patella tendon-bone (BTB) and hamstring autografts. Thin dots and lines: individual patients; thick line: modeled mean; dark grey ribbon: one standard deviation; light grey ribbon: two standard deviations.

**Figure 6.**
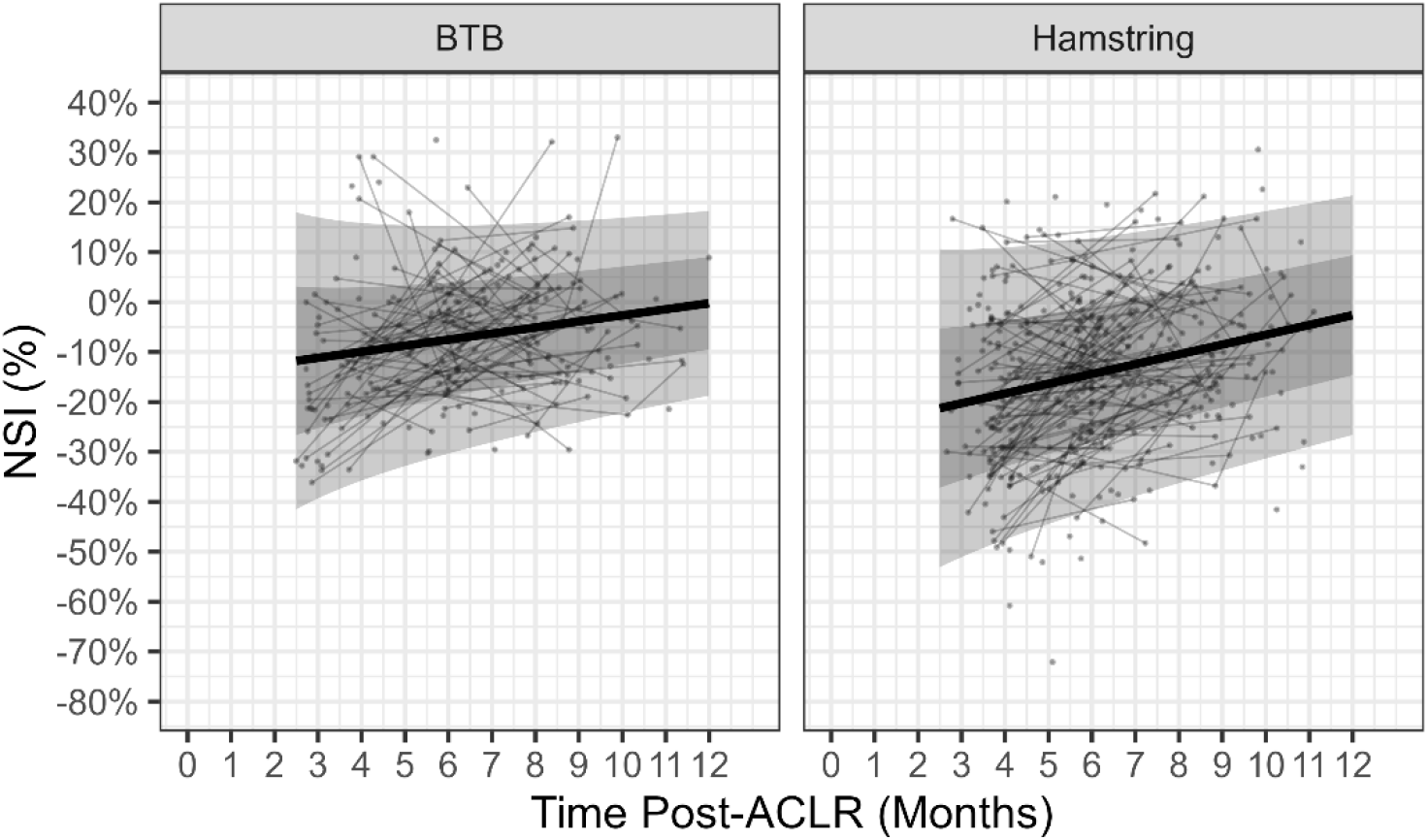
Normalize symmetry index (NSI) for knee flexion strength as a function of time post-anterior cruciate ligament reconstruction (ACLR) and autograft for bone-patella tendon-bone (BTB) and hamstring autografts. Thin dots and lines: individual patients; thick line: modeled mean; dark grey ribbon: one standard deviation; light grey ribbon: two standard deviations.

## DISCUSSION

This study provides insights into the dynamics of KE and KF muscle strength recovery following ACLR, highlighting the differential impacts of BTB and HAM autografts. Our hypothesis was supported as KE NSI increased and became more symmetrical over time post-ACLR, independent of autograft, and HAM patients exhibited more symmetrical KE NSI compared to BTB patients. Additionally, KE INV and KE UNINV increased with time post-ACLR and HAM patients displayed greater KE INV strength compared BTB patients, but there was no difference between autografts for KE UNINV. Together, this indicates that BTB autografts are associated with greater KE strength deficits which must be addressed during rehabilitation. In contrast, no differences were observed for either KF INV or KF UNINV between autografts. However, a significant difference was detected for KF NSI. This suggests that NSI may reveal muscle strength recovery disparities between autografts that are not apparent when examining the strength of INV and UNINV in isolation. Such findings emphasize the importance of evaluating both the KE and KF muscle strength of each limb independently as well as the difference between limbs to fully understand muscle strength recovery following ACLR. These findings align with existing literature, as substantial KE INV deficits in BTB patients have been noted ^4-6^ and highlights the importance of autograft in ACLR rehabilitation and emphasizes the need for tailored therapeutic strategies that consider autograft to optimize strength recovery.

This study delineates patterns of KE and KF muscle strength recovery over time post-ACLR, with attention to differences by autograft. By capturing how KE and KF muscle strength evolves during the first 12 months post-ACLR, these findings contribute to a clearer understanding of typical muscle strength recovery trajectories and reveal meaningful disparities in muscle strength recovery between autografts. This information can inform clinical expectations, support earlier identification of patients at risk for delayed muscle strength recovery, and guide more personalized rehabilitation strategies. While these findings provide valuable insight into the muscle strength recovery process, they also underscore the need for large-scale, population-level data to generate clinically useful normative reference values. Establishing such normative benchmarks would allow sports medicine providers to compare an individual patient’s muscle strength recovery progress against expected trajectories derived from broader patient populations, facilitating a more objective assessment of whether a patient is on track, ahead of, or lagging behind rehabilitation milestones. Furthermore, integrating muscle strength recovery trajectories with reference values from healthy, age-, sex-, and sport-matched individuals could enable a dual-layered assessment framework—evaluating both intra-individual muscle strength recovery and readiness for RTS based on strength parity with healthy peers. ^25^ This represents a logical next step toward enhancing decision-making in ACLR rehabilitation and RTS.

Ultimately, improving the ability to characterize KE and KF muscle strength recovery post-ACLR is critical as approximately 25% of young, high-demand ACLR patients experience reinjury of either the ACLR graft or the contralateral native ACL. Notably, nearly half of these re-injuries occur within the initial year following ACLR, highlighting potential deficiencies in the criteria used for RTS testing that, in part, aim to help sports medicine providers determine when modifiable risk factors are adequately addressed and the risk of re-injury is acceptably reduced.^26-29^ Thigh muscle weakness is commonly believed to be one of these modifiable risk factors ^30^; however, despite apparent support for RTS testing, 41% of published studies have implemented strength testing as a criteria used to clear patients to RTS.^15^

When objective criteria are used, the most frequent metric reported is KE NSI or a similarly computed symmetry index.^15,18^ In numerous RTS tests, a patient is required to achieve an NSI that demonstrates less than a 10% deficit in INV compared to UNINV, which is assumed to indicate symmetrical strength or resolved INV weakness, before being ‘cleared’ to RTS.^15^

Despite the strong focus on the KE NSI and the intuitive advantage of having symmetrical rather than asymmetrical strength, evidence linking symmetrical strength directly to a reduced risk of ACL re-injury remains scarce.^30^ This could be because a patient may not have symmetrical strength prior to injury and bilateral strength deficits can occur after ACLR, suggesting that using UNINV as a control to indicate resolved INV deficits might not be appropriate.^31-33^ Further, it is possible that many individuals may exhibit what appears to be symmetrical strength, yet this symmetry often masks an underlying bilateral weakness that would not be recognized if KE NSI were used in isolation to ‘clear’ patients for RTS. ^34^ Nevertheless, despite the emphasis placed on the 10% deficit or less threshold, a low percentage of patients are noted to achieve this at the time of RTS regardless of autograft ^35-37^, which was corroborated by the present study. This may be because KE INV and KE UNINV muscle strength both improve over time post-ACLR, perhaps indicating that using KE UNINV as a reference to compare KE INV to is a ‘moving target’ ^38^ resulting in overestimation of function. ^31^ Nonetheless, while NSI has its limitations, it still offers valuable insights into the laterality of muscle strength when integrated thoughtfully within the broader framework, aiding in more holistic RTS evaluations.

There are some limitations of this retrospective study. This study analyzed data collected during routine clinical practice, which presents several challenges. The data were irregularly spaced as patients underwent varying numbers of RTS tests at different intervals. Specifically, RTS tests were more frequent between 4 to 8 months post-ACLR, resulting in a non-uniform distribution over time, with most patients having two or fewer RTS test. Although the utilized statistical methodology is resilient to these issues, a more optimal approach would have involved standardized collection of KE and KF muscle strength data at fixed intervals, such as at 3, 6, 9, and 12 months post-ACLR, to achieve a more even distribution and greater density of observations per patient. This would also allow for the modeling of non-linear effects of time post-ACLR, which may be present as KE and KF muscle strength recovery likely display greater rates of recovery between 3 and 9 months post-ACLR and then begin to plateau. Although, data collection ceased at 12 months post-ACLR, hence longer-term recovery trends remain unexplored. This study’s cohort was limited to ACLR patients aged 12 to 30 years; therefore, these results may not be generalizable to all age groups, despite this age range being most susceptible to ACL injuries. While all participants were involved in recreational activities or sports pre-injury as reflected by the mean pre-injury MARX score, the study did not measure competition level, RTS desires and motivation, or long-term RTS status, factors that could influence rehabilitation engagement and muscle strength recovery outcomes. Finally, even though we controlled for the effect of concurrent meniscal and articular cartilage procedures, additional surgical details were not considered in the analysis, which may influence muscle strength outcomes; although, this could be considered a strength as our data reflects a high number of surgeons, potentially enhancing the generalizability of our findings.

In conclusion, this study delineates the nuanced impacts of autograft on the trajectory of KE and KF muscle strength recovery following ACLR. The findings underscore the superior recovery of KE muscle strength in HAM patients compared to BTB patients, highlighting the influence of autograft on rehabilitation outcomes. These insights enhance our understanding of post-ACLR strength muscle strength recovery dynamics and support more informed clinical decision-making regarding patient monitoring and rehabilitation planning. The development of population-level normative recovery trajectories—stratified by autograft type—could further refine the ability to assess patient progress, personalize rehabilitation, and optimize timing for RTS decisions.

## Data Availability

Data produced in the present study are available upon reasonable request to the authors.

